# Systematic analysis and prediction of genes associated with disorders on chromosome X

**DOI:** 10.1101/2022.02.16.22270779

**Authors:** Elsa Leitão, Christopher Schröder, Ilaria Parenti, Carine Dalle, Agnès Rastetter, Theresa Kühnel, Alma Kuechler, Sabine Kaya, Bénédicte Gérard, Elise Schaefer, Caroline Nava, Nathalie Drouot, Camille Engel, Juliette Piard, Bénédicte Duban-Bedu, Laurent Villard, Alexander P.A. Stegmann, Els K. Vanhoutte, Job A.J Verdonshot, Frank J. Kaiser, Frédéric Tran Mau-Them, Marcello Scala, Pasquale Striano, Suzanna G.M. Frints, Emanuela Argilli, Elliott H. Sherr, Fikret Elder, Julien Buratti, Boris Keren, Cyril Mignot, Delphine Héron, Jean-Louis Mandel, Jozef Gecz, Vera M. Kalscheuer, Bernhard Horsthemke, Amélie Piton, Christel Depienne

## Abstract

Disease gene discovery on chromosome (chr) X is challenging owing to its unique modes of inheritance. We undertook a systematic analysis of human chrX genes. We observe a higher proportion of disorder-associated genes and an enrichment of genes involved in cognition, language, and seizures on chrX compared to autosomes. We analyze gene constraints, exon and promoter conservation, expression and paralogues, and report 127 genes sharing one or more attributes with known chrX disorder genes. Using a neural network trained to distinguish disease-associated from dispensable genes, we classify 235 genes, including 121 of the 127, as having high probability of being disease-associated. We provide evidence of an excess of variants in predicted genes in existing databases. Finally, we report damaging variants in *CDK16* and *TRPC5* in patients with intellectual disability or autism spectrum disorders. This study predicts large-scale gene-disease associations that could be used for prioritization of X-linked pathogenic variants.

## Introduction

Sex in mammals is determined by a diverging pair of sex chromosomes (chr). Human females have two copies of the 155-Mb chrX while males have a single X copy and a smaller 30-Mb chrY. Compensation of gene dosage in females is achieved through X chromosome inactivation (XCI), a process leading to the epigenetic silencing of an entire chrX, apart from two pseudoautosomal regions (PARs). This process happens during early embryonic development, is randomly and independently established in each cell, and stably maintained during further cell divisions.^1,2^ As a consequence, female individuals are cell mosaics, each cell expressing genes from either the maternal or paternal X copy.^3^ A subset of genes, which can be variable between individuals and tissues, escapes X inactivation and continues to be expressed from both X chromosomes.^4^

The last two decades have revolutionized concepts of X-linked inheritance, by depicting its unique but multiple forms.^1,5^ The first and most widely described disorders (>100 genes) mainly affect hemizygous males and are transmitted through healthy or mildly symptomatic female carriers. Other modes of X-linked inheritance are mainly observed in disorders affecting the central nervous system. Variants in X-linked genes such as *MECP2*^6^, *CDKL5*^7,8^ and *DDX3X*^9^ preferentially affect heterozygous females. These variants usually occur *de novo*, and hemizygous males are either not viable or survive only if variants are hypomorphic or mosaic. Variants in other genes affect hemizygous males and heterozygous females almost equally. The list of X-linked disorders first described as selectively affecting males but turning out to also affect females is continually increasing and include *IQSEC2*^10^, *NEXMIF*,^11^ *KDM5C*,^12^ *HUWE1*,^13^ *USP9X*^14^ and *CLCN4.*^15^ Lastly, two X-linked disorders, related to *PCDH19* and *EFNB1*, affect heterozygous females and postzygotic somatic mosaic males (due to cellular interference), while hemizygous males are spared.^16,17^

Disease gene discovery on chrX is thus associated with greater challenges, including male-female patient selection and variant interpretation biases, compared to autosomes. ChrX is often omitted from genome-wide analyses in a research context due to the difficulty of dealing with sex dichotomy in bioinformatics pipelines. Identifying novel gene-disease associations on chromosome X requires dedicated studies of families with multiple affected males^18,19^ or multiple subjects with matching phenotypes.^9^ The interpretation of X-linked variants in sporadic cases and small families remains difficult in absence of extended segregation in the family, which is rarely available. Furthermore, the presence of damaging variants at relatively high frequency in large databases (ExAC, gnomAD) led to question previously established gene-disease associations.^20^ The Deciphering Developmental Disorders (DDD) consortium recently estimated that X-linked disorders overall affect males and females equally and represent 6% of developmental disorders.^21^ However, despite the remarkable size of the cohort (11,044 affected individuals), this study failed to identify new X-linked disorder-associated genes.

In this study, we first undertook a systematic analysis of all coding genes on human chromosome X and compared the proportion and characteristics of associated disorders to those on autosomes. In a second step, we investigated the relevance of multiple variables to predict gene-disorder associations. Lastly, we used these predictions to uncover new disease-gene associations supported by the literature as well as by patient data and functional studies.

## Results

### Chromosome X is enriched in disorder genes and in genes relevant to brain function

Chromosome X comprises 829 protein coding genes annotated in HUGO Gene Nomenclature Committee (HGNC), including 205 associated with at least one monogenic disorder (referred to as ‘disorder genes’) in OMIM (Fig. 1a). We used the clinical synopsis to compare the proportion of disorder genes and their associated clinical features on chrX (available for 202 genes) and autosomes (Supplementary Methods, Fig. 1b). We observed a significant and specific enrichment in disorder genes on chrX (24% versus 12-22%, *p*=1.87×10^-3^; OR=1.5; Fig. 1c-d and Supplementary Tables 1-4). Furthermore, genes on chrX were significantly more frequently associated with neurological phenotypes than genes on autosomes (77% versus 55-76%; *p*=7.01×10^-3^; OR=2.0; Fig. 1e-f). More specifically, we observed that chrX is enriched in genes associated with intellectual disability (ID; 58% versus 27-45%; *p*=5.92×10^-11^; OR=2.9), seizures (46% versus 23-38%; *p*=1.12×10^-4^; OR=2.1) and language impairment (32% versus 11-24%; *p*=5.59×10^-3^; OR=2.0; Fig. 1g-l), but not motor development, spasticity, or ataxia (Fig. 1m-n; Extended Data Fig. 1a-b; Supplementary Table 5). The difference remained significant when genes associated with provisional gene-phenotype relationships (P), susceptibility to multifactorial disorders (M) or traits (T) (referred to as ‘PMT’ genes) were included in the comparison (Extended Data Fig. 1c-d; Supplementary Tables 4 and 5). In total, 84% of known disease genes on chrX are associated with ID, seizures or language impairment and about 30% are associated with all three clinical outcomes (Fig. 1o).

**Fig. 1:**
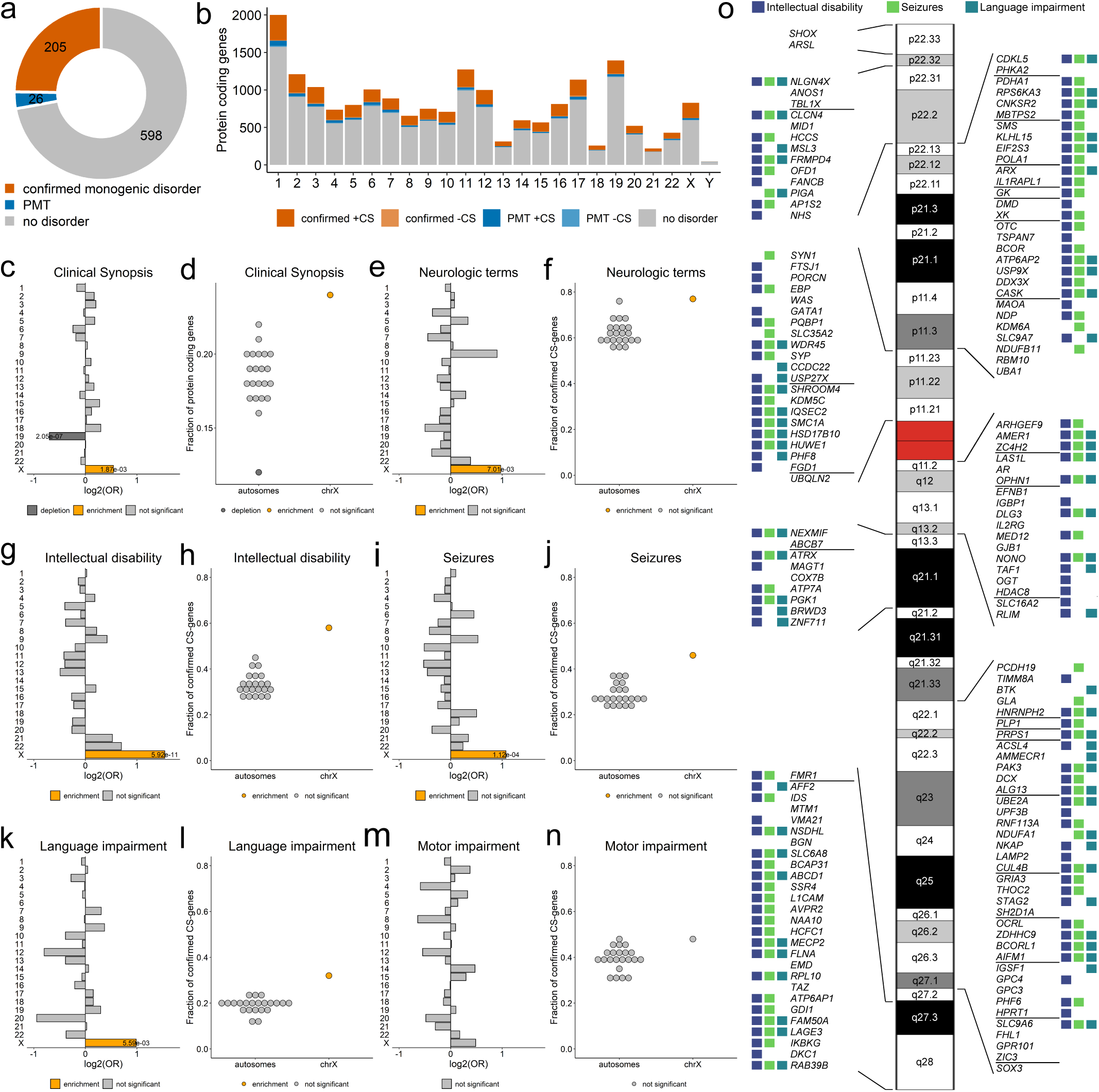
Disorder genes across chromosomes. **a,** Number of protein-coding genes on chrX. Genes associated with at least one monogenic disorder: orange, confirmed genes; blue: genes with provisional associations (P), associated with susceptibility factors to multifactorial disorders (M) or traits (T) (altogether: PMTs); grey: genes without known phenotypes (no-disorder genes). **b,** Number of protein-coding genes *per* chromosome. Dark and light colors represent genes with Clinical Synopsis (CS) data for at least one of the associated phenotypes (+CS) or without CS (-CS), respectively. **c,** *Per* chromosome enrichment/depletion of protein-coding genes with at least one associated phenotype comprising Clinical Synopsis data (confirmed CS-genes). **d,** Fraction of confirmed CS-genes among protein-coding genes. **e-n.** Predominance of chrX genes associated with neurologic features. **e, g, i, k and m,** *Per* chromosome enrichment/depletion of genes with non-specific neurologic features (**e**), intellectual disability (**g**), seizures (**i**), language impairment (**k**) or motor development (**m**). **f, h, j, l and n,** Fraction of genes associated with non-specific neurologic features (**f**), intellectual disability (**h**), seizures (**j**), language impairment (**l**) or motor development (**n**). **c-n**, Yellow, enrichment; darkgrey, depletion; light grey, not significant. Terms corresponding to the same neurological clinical features were used in OMIM searches (Supplementary Table 2). Only significant p-values are shown (Fisher’s test followed by Bonferroni correction for multiple testing; Supplementary Tables 4 and 5). **o,** Scheme of genes associated with neurological features on chrX. Squares next to the genes represent association with intellectual disability (blue), seizures (green) or language impairment (cyan). A horizontal line separates genes present in different chromosome bands.

### Confirmed disorder-associated genes share specific features

Despite chrX being enriched in disorder-associated genes, 598 genes (71%) are not yet related to any clinical phenotype (referred to as ‘no-disorder genes’; Fig. 1a). We hypothesized that disorder genes share specific common features that dispensable genes do not exhibit and that could be used to predict genes that remain to be associated with human disorders. To test this hypothesis, we retrieved annotations from different sources and/or calculated additional metrics, including: 1) gnomAD gene constraint metrics: LOEUF (rank of intolerance to loss-of-function (LoF) variants), misZ (intolerance to missense variants score) and synZ (intolerance to synonymous variants score); 2) coding-sequence (CDS) length; 3) the degree of exons and promoter conservation across 100 species; 4) the promoter CpG density;^22^ and 5) gene expression data, conveyed as a tissue specificity measure (tau) and brain-related expression levels from GTEx and BrainSpan resources (Supplementary Table 1).

We investigated whether the distribution of these variables differ between disorder genes and no-disorder genes. As expected, disorder genes had lower LOEUF values and higher misZ scores but similar synZ scores than no-disorder genes. We observed a significant enrichment of disorder genes in the three lowest LOEUF deciles and in the two highest misZ deciles, with the distributions of LOEUF and misZ metrics in the PMT group showing intermediate values between disorder and no-disorder genes (Fig. 2a-b). Disorder genes were enriched in the highest decile of exon conservation, promoter conservation scores and CDS length distribution with an overall distribution that was variable in all groups (Fig. 2c; Extended Data Fig. 2; Supplementary Table 6). Disorder genes tended to present higher promoter CpG density than no-disorder genes, although the difference was not significant. Remarkably, we observed that disorder genes are more broadly expressed (tau<0.6) and show intermediary levels of gene expression in brain-tissues compared to no-disorder genes, for which expression was more often restricted to a few tissues (tau>0.6) and are less expressed in the brain (Extended Data Fig. 2).

**Fig. 2:**
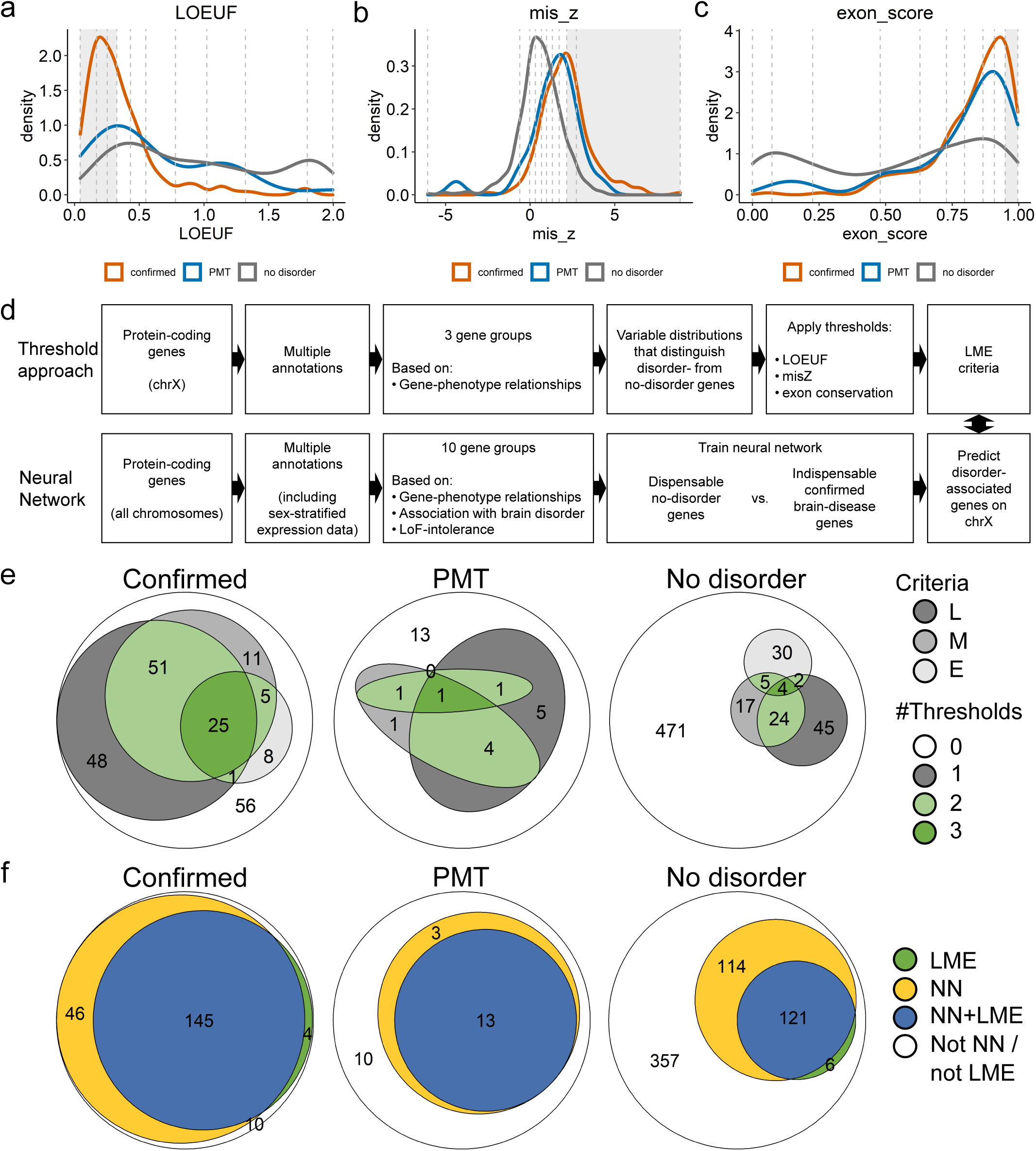
Prediction of disorder-genes. **a-c,** Density plots showing the distribution of LOEUF (**a**), misZ (**b**) and exon-conservation score (**c**) according to gene group. Confirmed disorder genes (orange), PMT genes (blue), no-disorder genes (grey). Vertical dashed lines separate deciles of the overall distribution. Grey areas depict deciles for which confirmed disorder-associated genes are enriched (related to Extended Data Fig. 2). **d,** Overview of the approaches used to uncover features shared by disorder genes and to predict new putative disorder-genes (details in Supplementary Methods). **e,**Euler diagrams showing the number of genes fulfilling LOEUF (L), misZ (M) and exon-conservation (E) criteria for confirmed (left), PMT (middle) and no-disorder genes (right). Thresholds: LOEUF ≤ 0.326, misZ ≥ 2.16, exon-conservation score ≥ 0.9491. Genes fulfilling at least two LME criteria are shown in green (light or dark green for two or three criteria, respectively). Genes meeting only one of the metrics are shown in different shades of grey (dark to light: LOEUF, misZ, exon-conservation). **f,** Euler diagrams showing the number of genes fulfilling LME criteria (LME, green), predicted by the neural network (yellow) or both (blue) for confirmed (left), PMT (middle) and no-disorder genes (right). **e and f,** Genes neither predicted by the neural network nor meeting LME criteria are shown in white.

We next investigated how the existence of close paralogues and the location in PARs may influence their relationship to human disease. From the 18 PAR protein-coding genes, only two (*SHOX* and *CSF2RA*) have been associated with a medical condition so far. We also observed that disorder genes are significantly depleted in close paralogues compared to PMT/no-disorder genes (*p*=3.1×10^-6^; OR = 0.33; Extended Data Fig. 3a-d). This suggests that highly similar paralogues might compensate pathogenic variants in corresponding X-linked genes.

Previous comparisons suggest that LOEUF, misZ, and exon conservation scores, which showed marked differences between the two subgroups, can best differentiate disorder from no-disorder genes. We then focused on the deciles enriched in disorder genes to list the genes in the no-disorder group exhibiting similar characteristics (Fig. 2d). More specifically, we defined thresholds for LOEUF, misZ, and exon conservation scores as follows: LOEUF ≤ 0.326 (L), misZ ≥ 2.16 (M), and/or exon-score ≥ 0.9491 (E). 149 of the 205 (73%) disorder genes met one or more L, M or E (LME) criteria (Fig. 2e). Among the 205 disorder genes, 56 genes failing to meet any of the LOEUF/misZ criteria had a smaller CDS length (*p*=4.20×10^-9^), indicating that the performance of LOEUF and misZ in differentiating disorder from no-disorder genes depends on CDS length (Extended Data Fig. 3e). Exon conservation alone allowed retrieving eight disorder genes with small CDS for which LOEUF and misZ failed to reach the thresholds (Extended Data Fig. 3e; Supplementary Table 1). When applying the same thresholds to the 598 genes not associated with disease, 127 (21%) genes fulfilled at least one condition and 35 fulfilled at least two criteria (Fig. 2e; Supplementary Table 1). Half (n=13) of the PMT genes also met at least one criterion and seven at least two criteria (Fig. 2e). Altogether, 140 genes shared at least one of the LME criteria with known disorder genes.

### Prediction of novel disorder-associated genes on chrX

The threshold approach was limited by the number of variables that could be taken into account to differentiate disorder from no-disorder genes. We then used machine learning to predict remaining disorder genes in a more systematic and unbiased fashion. As most X-linked disorders are associated with neurological features, we trained a neural network (NN) to distinguish two classes of genes: 1) known disorder genes associated with neurological features on all chromosomes and 2) dispensable genes as defined by Karczewski et al. (2020)^23^ (Supplementary Methods; Extended Data Fig. 4; Extended Data Fig. 5a-b). We used additional variables including expression data stratified according to sex and data on paralogues (Supplementary Tables 7 and 8). Although using data on all chromosomes, the neural network performed remarkably better for genes on chrX, predicting 191 (93%) of the 205 known disorder genes on this chromosome using FDR < 0.05 (Extended Data Fig. 5c-f). 145 of the predicted genes met the LME thresholds (Fig. 2f). The NN confirmed 16 of the 26 PMT genes as disease-associated, 13 of which also met LME criteria (Fig. 2f, Fig. 3a-b). Focusing on the 598 no-disorder genes on chrX, the NN predicted 235 as disease-associated (FDR < 0.05; Supplementary Table 9; Extended data Fig. 5a). 121 genes (51%) also met at least one of the LME criteria (Fig. 2f), with four meeting all three, 30 two and 87 genes one criterion (Fig. 3c). The 114 remaining genes predicted by the NN did not meet any LME criteria (Fig. 3d), whereas only six meeting LME criteria were not predicted by the NN (Fig. 3e). Altogether, these results suggest that the NN is a valid approach to predict genes remaining to be associated with disease with high accuracy.

**Fig. 3:**
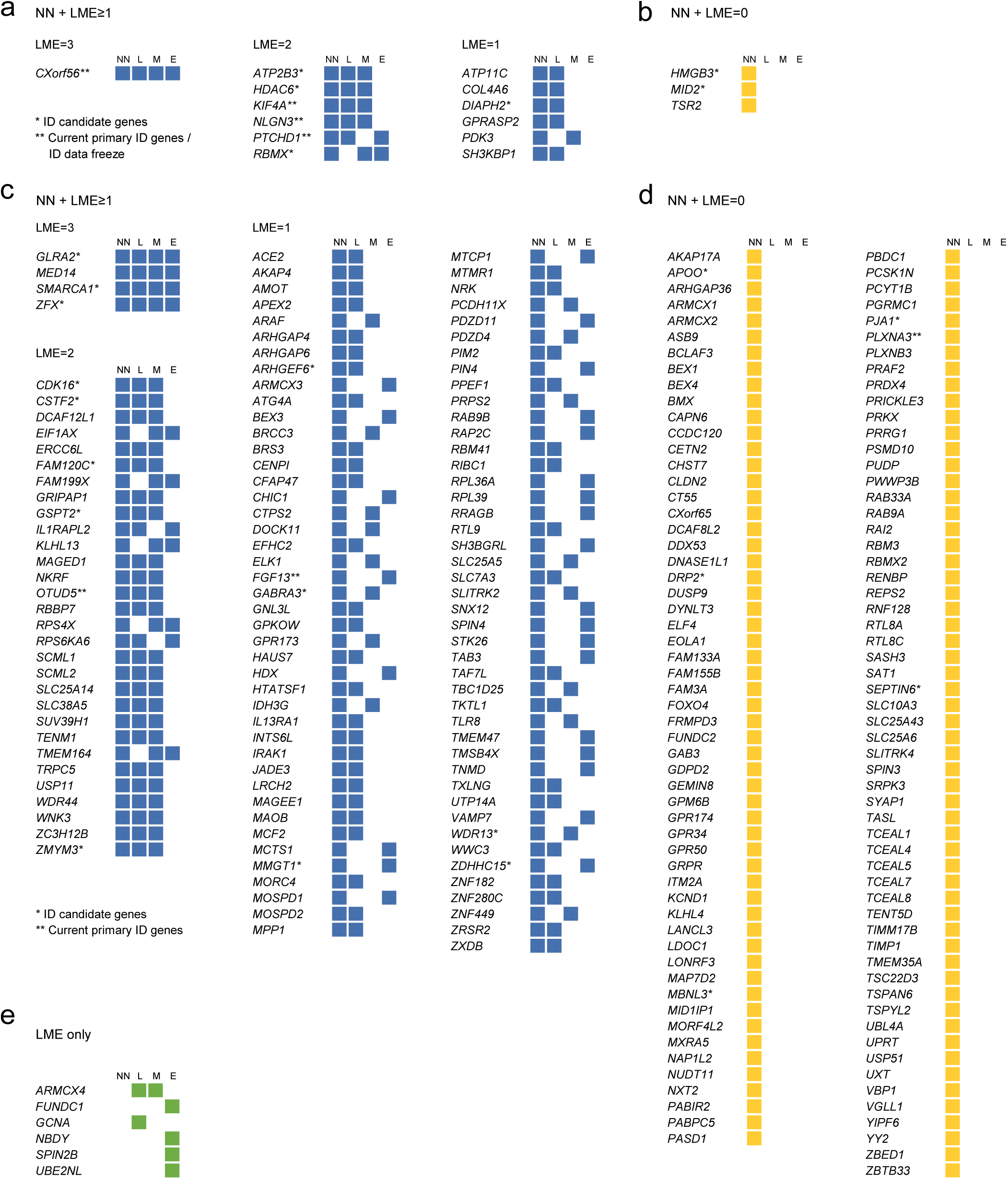
Different evidence strength for the predicted disorder-associated genes. **a-d,** PMT (**a, b**) and no-disorder genes (**c, d**) predicted by the neural network as possibly disease-associated, either fulfilling (**a, c**) or not LME criteria (**b, d**). Genes present in the SysID database as “ID candidate genes” (*) and “Current primary ID” or “ID data freeze” (**) are marked. **e,** No-disorder genes meeting LME criteria but not predicted by the neural network.

### Validation of putative novel disorder genes

To provide additional evidence supporting our predictions, we searched for evidence from databases or the literature that variants in the 235 NN-predicted genes could indeed result in new X-linked disorders. We first used HGMD and DECIPHER to retrieve the number of single nucleotide variants and indels of unknown significance reported in each gene and compared the number of variants in predicted versus non-predicted gene categories (Supplementary Table 9). This analysis revealed that the 235 NN-predicted disorder genes are enriched in point variants compared to non-predicted genes (*p*=5.3×10^-6^; OR=2.2), suggesting that this excess is due to pathogenic variants. NN-predicted genes meeting at least one LME criterion had more point variants than predicted genes that do not meet any LME criterion (Fig. 4a-b; Extended Data Fig. 6).

**Fig. 4:**
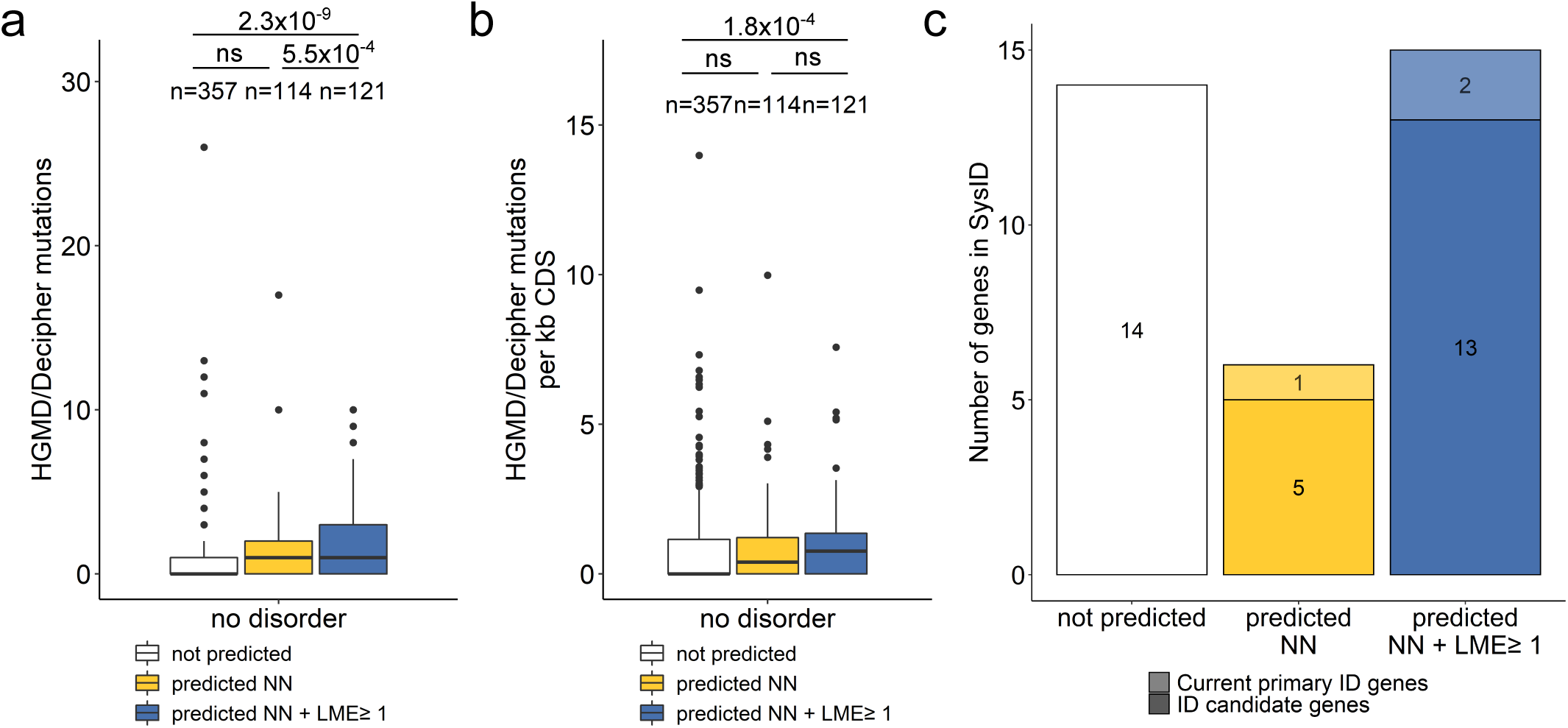
External supporting evidence for the predicted disorder-associated genes. **a and b,** Known point mutations in no-disorder genes. Boxplot showing the number of known mutations reported in HGMD and DECIPHER (**a**) or their value normalized by coding-sequence (CDS) length (**b**) according to their predicted status. No-disorder genes meeting LME criteria but not predicted by the neural network were omitted from the analysis. Box plot elements are defined as follows: center line: median; box limits: upper and lower quartiles; whiskers: 1.5× interquartile range; points: outliers. Related to Supplementary Table 9. **c,** Number of nodisorder genes present in the SysID database according to their predicted status.

Second, we used the SysID and Gene2Phenotype (G2P) developmental disorders databases to compare the overlap of predicted and non-predicted genes (Fig. 4c). Nine of the 16 NN-predicted PMT genes were in SysID (Fig. 3a-b). Three genes (*PTCHD1*, *NLGN3*, and *KIF4A*), indicated as confirmed SysID genes, were also in G2P. *PTCHD1* and *NLGN3* are associated with susceptibility to autism in OMIM, but both were recently confirmed to cause a monogenic ID disorder frequently associated with autism.^24–26^ A splice site variant in *KIF4A* was identified in a family with four affected males,^27^ and this finding was recently strengthened by the identification of additional *de novo* or inherited variants causing a range of different phenotypic manifestations.^28,29^ Focusing on no-disorder genes, 21 (9%) of the NN-predicted genes and 14 (4%) of the non-predicted genes were in SysID (Fig. 3c-d). Five predicted genes (*CSTF2*, *FGF1*3, *PLXNA3*, *OTUD5* and *STEEP1*) were confirmed SysID genes. In addition, six genes (*ARHGEF6*, *CDK16*, *FGF13*, *GSPT2*, *MMGT1* and *ZDHHC15)* had entries in G2P. Pathogenic variants in *OTUD5* and *FGF13* respectively cause a severe neuro-developmental disorder with multiple congenital anomalies and early lethality,^30,31^ and developmental and epileptic encephalopathy,^32^ both described in 2021. *PLXNA3* have been associated with ID and autism spectrum disorders (ASD),^33^ or hypogonadotropic hypogonadism.^34^ Additional evidence from the literature not yet reflected in OMIM or any other database include possibly pathogenic variants described in one or a few families in *ZMYM3*,^35^ *GLRA2*,^36^ and *GPKOW.*^37^ Although some of these genes now have an OMIM disorder entry, their association with a monogenic disorder was not known at the beginning of our study.

Third, we retrieved the number of *de novo* predicted damaging (truncating or CADD≥25) variants identified in these genes from the Martin et al. ^21^ and Kaplanis et al.^38^ studies. In parallel, we examined exome data from two additional cohorts of patients with developmental disorders, containing mainly patients with intellectual disability (6500 from 2346 families [Paris-APHP cohort] and 1399 individuals from 463 families [UCSF cohort) and extracted predicted damaging variants in NN-predicted genes meeting at least one LME criterion (Supplementary Table 10). This led us to select 13 genes containing *de novo* damaging variants and additional predicted damaging variants identified in independent cohorts (Fig. 5). Interestingly, 19 out the 55 missense variants reported in these 13 genes are localized in known functional protein domains (Fig. 5; Supplementary Table 11). Among those genes, *SMARCA1* has been associated with syndromic intellectual disability and Coffin-Siris-like features by the Clinical Genome Resource (ClinGen) with moderate evidence.^39^

**Fig. 5:**
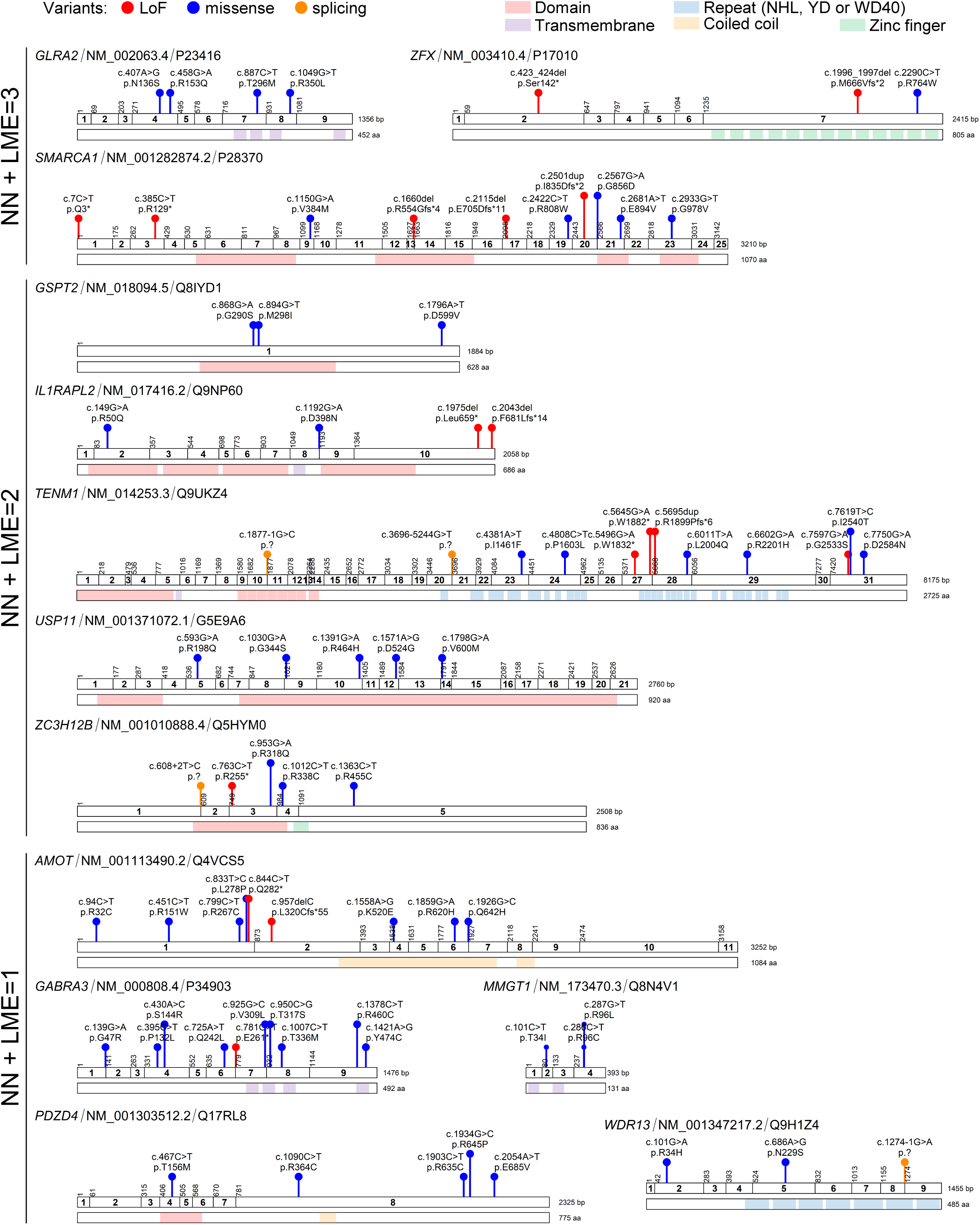
Damaging variants in selected predicted disorder-associated genes. Schematic representation of the coding exons, protein domains (when present) and available damaging variants (truncating or CADD≥25) for each selected predicted gene. Types of variants are shown in different colors: lof-of-function (LoF, red), missense (blue), splicing (orange). Protein functional domains are shown: domains (light red), transmembrane segments (purple), NHL, YD or WD40 repeats (light blue), coiled-coils (yellow), zinc-fingers (green). HGVS cDNA and HGVS protein descriptions are shown. The corresponding RefSeq identifier of the MANE Select transcript and Uniprot identifier are shown for each gene. Details of variants displayed in this figure appear in Supplementary Table 11.

### *CDK16* and *TRPC5* are novel genes associated with intellectual disability

Lastly, we focused on *CDK16* and *TRPC5*, both predicted by the NN and threshold approaches but for which genetic evidence was limited (Fig. 6a-b). *CDK16* encodes a protein kinase involved in neurite outgrowth, vesicle trafficking and cell proliferation.^40^ A deletion of two nucleotides leading to a frameshift (NM_006201.5: c.976_977del, p.(Trp326Valfs*5)) segregating in four males with ID, ASD, absence seizures and mild spasticity was reported in this gene by Hu et al. ^18^ Using exome sequencing, we identified a nonsense variant (c.961G>T, p.(Glu321*)) in an adult patient with ID and spasticity. A missense variant (c.1039G>T, p.(Gly347Cys)) affecting a highly conserved amino acid of the kinase domain (CADD PHRED score: 32) was identified by genome sequencing in a male patient with ID, ASD and epilepsy, whose family history was compatible with X-linked inheritance (Fig. 6a and c; Supplementary Table 12). In addition, a nonsense variant (c.46C>T, p.(Arg16*)) was recently reported in a patient with ASD by Satterstrom et al.^41^

**Fig. 6:**
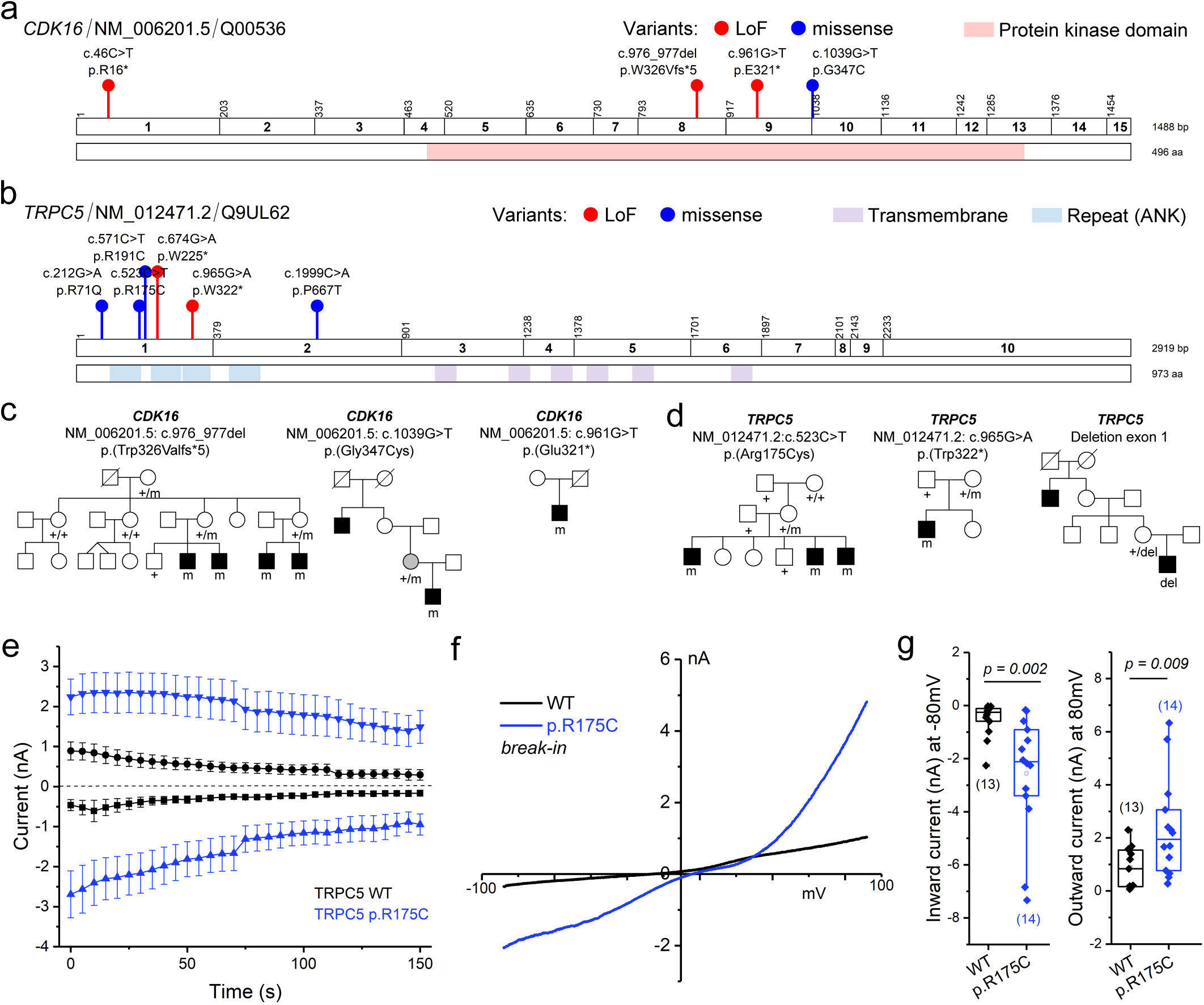
Validation of selected predicted disorder-associated genes: *CDK16* and *TRPC5*. **a and b,** Schematic representation of the coding exons, protein domains and variants of *CDK16* (**a**) and *TRPC5* (**b**). Variant types: loss-of-function (LoF, red), missense (blue). **c,** Pedigrees of the three families with *CDK16* variants. The pedigree of family with c.976_977del, p.(Trp326Valfs*5) was adapted from Hu et al. (2016)^18^. The two remaining families are unpublished. **d,** Pedigrees of the three families with *TRPC5* variants. The family with an intragenic exon deletion was reported in Mignon-Ravix et al. (2014)^43^. The two other families are novel. **e**-**g,** Functional characterization of *TRPC5* p.R175C by whole-cell patch-clamp recordings showing that the mutation renders the mutant channel constitutively opened. **e,** Time course of inward and outward current amplitudes measured at +80 mV and -80 mV in HEK293 cells transiently expressing WT (black) and mutant TRPC5 (red) in presence of 100nM free Ca^2+^ in the pipette. Values are reported as mean ± SEM. Recordings started few seconds after the rupture. **f,** Representative whole-cell current-voltage (I-V) relationships of WT and mutant TRPC5 channels current obtained shortly after break-in (≤ 10 s) with 100nM free Ca^2+^ in the pipette. **g,**Boxplot of WT and mutant whole-cell current at -80 mV and +80 mV after break-in (one-way ANOVA with Dunnett’s post hoc test). The number of independent recordings appears in brackets. Details of variants displayed in this figure appear in Supplementary Table 12.

*TRPC5* encodes the short transient receptor potential channel 5, a channel permeable to calcium predominantly expressed in the brain.^42^ We identified a missense variant in this gene (NM_012471.2:c.523C>T, p.(Arg175Cys), CADD PHRED score: 29.8) in three male siblings with ID and ASD. The variant was maternally inherited but absent from the maternal grandparents (Fig. 6b and d; Supplementary Table 12). We investigated the functional impact of the p.(Arg175Cys) variant on the TRPC5 channel using whole-cell patch-clamp. Immediately after break-in, HEK293 cells expressing mutant TRPC5 exhibited an increase in immediate current recorded compared with cell expressing wild-type TRPC5 (p= 0.009 for outward current at -80mV and p=0.002 for inward current at -80mV; Fig.6 e-g), suggesting a constitutively active current. A nonsense variant (c.965G>A, p.(Trp322*)) was identified by exome sequencing in a patient with high-functioning ASD. An intragenic deletion of the first coding exon of *TRPC5*, encoding conserved ankyrin repeats, was previously reported in a patient with ASD who had a family history compatible with X-linked inheritance.^43^ In addition, three *de novo* variants in *TRPC5* (p.(Pro667Thr), p.(Arg71Gln), p.(Trp225*)) had been identified in patients with intellectual disability and/or autism disorders in the literature.^21,44,45^

Altogether, these results strongly suggest that pathogenic variants altering the functions of *CDK16* and *TRPC5* lead to novel X-linked disorders featuring ID and ASD.

## Discussion

Male and female individuals tolerate pathogenic variants on chrX in different manners. Variants in X-linked genes have to be interpreted taking this complexity into account. We aimed at providing an inventory of disorder genes on chrX and predict genes that remain to be associated with human disease, assuming that they share similar characteristics. We first used a threshold approach to list genes similarly constrained during evolution, which are the most likely to lead to disease when altered by genetic variants. This revealed 127 novel genes sharing at least one constraint metric with known disorder genes and 35 sharing at least two. To avoid bias and limitations linked to this approach, we used a hypothesis-free neural network to differentiate genes associated with brain disorders from genes where damaging variants are tolerated (dispensable genes). The NN predicted 235 genes as putative disorder genes, including most of the genes uncovered by the threshold approach.

Our predictions are supported by a higher number of point variants reported in DECIPHER and HGMD in predicted versus non-predicted genes, which strengthens the probability that variants reported contribute to the patients’ phenotypes and prompts the prioritization of these genes in further genetic analyses. We notably highlight 13 genes in which several possibly pathogenic variants in functional domains have been identified in patients, likely constituting novel neurodevelopmental disorder genes. Furthermore, a subset of predicted genes (e.g. *OTUD5*, *FGF13*, *GLRA2* and *GPKOW)* have already been associated with human diseases, although these associations had no OMIM Gene-Phenotype Relationship entries when we started our study. Furthermore, we provide additional evidence that variants in two predicted genes, *CDK16* and *TRPC5*, likely cause novel X-linked disorders by gathering genetic and clinical data of unrelated families with damaging variants in these genes.

Altogether, our gene-focused approach therefore suggests that less than half of genes associated with human pathology on chrX are known so far and many more remain to be characterized clinically, which challenges the recent estimation that X-chromosome has been saturated for disease genes.^21^ The differences in conclusions may come from recruitment bias in the DDD study, with inclusion of families with X-linked inheritance in more specific studies, as well as from the patient-driven approach itself, which only allows detecting what is statistically significant in a sample, no matter how large this sample is. It is indeed possible or even likely that phenotypes associated with genes we predict are ultra-rare, lethal *in utero*, difficult to recognize or assess clinically, or associated with atypical modes of X-linked inheritance, making their identification difficult using classical genetic approaches.

Focusing on known genes associated with monogenic conditions in the first part of this study, we observed a higher proportion of genes associated with disorders on chrX compared to autosomes. This finding suggests that family-driven approaches have been more efficient in identifying disorder-associated genes on this chromosome. Nevertheless, considering disease-causing genes independently of their proportion, we also observed an enrichment of genes associated with specific neurological features such as ID, seizures, and language delay on chrX. The specific association of X-linked genes with cognitive functions has largely been conveyed in the literature but, to our knowledge, this study is the first demonstrating the statistical significance of this finding using a systematic unbiased analysis. The reason why so many genes important for cognition and language are on a sex chromosome is fascinating but remains so far mysterious.^46^

Our study also indicates that disorder genes on chrX are depleted in highly similar paralogues (>95% identity) compared to PMT/no-disorder genes, a finding that remained significant for predicted genes compared to non-predicted ones (p<2.2×10^-16^; OR= 0.22). The consequence of the existence of paralogues for disorders is probably different depending on the expression of these paralogues and the ability of the gene product to compensate the function of the original genes. Interestingly, 20% of genes on chrX have paralogues with >95% identity. This list includes copies or retrocopies on autosomes, a functional redundancy that has been attributed at least partly to the transcriptional silencing of chromosome X during spermatogenesis.^47,48^ Although many gene copies or retrocopies are specifically expressed in male germ cells, others are still broadly expressed and could therefore buffer genetic variants in the X-linked paralogous genes, as shown for *UPF3B* and *UPF3A*.^49^ The existence of paralogues is also linked in some ways to the location in PAR regions and escape to XCI. Indeed, genes in PAR have paralogues on chrY and escape inactivation.^50^ Accordingly, 35 of the 231 predicted genes outside PAR also show some degree of escape from X inactivation (Supplementary Table 1). Interestingly, genes escaping XCI outside PAR, including *IQSEC2* and *KDM5C*, may lead to disorders manifesting in both sexes.^10,12^ Our study indicates that genes in PAR and genes with close paralogues are less likely to be associated with a disorder, suggesting that paralogues can compensate pathogenic variants in some X-linked genes and raising the possibility of digenism or oligogenism in genes with redundant functions.

Our study focused on coding genes and did not include genes encoding long non-coding RNA (lncRNA) or other RNA classes, which are however very abundant on chrX. Only a few non-coding genes have been associated with disease so far and we believe that constraints and pathological mechanisms applying to non-coding genes are likely different from those of coding genes. In this respect, our study only predicts genes associated with disorders when affected by usual mutation types. Therefore, our findings do not exclude that non-predicted genes could lead to disease when associated with unusual mechanisms, such as gain of a new function, dominant-negative impact on another gene, and ectopic expression of a gene in the wrong tissue or at an abnormal time during development.

In conclusion, our study provides new insights into the complexity of X-linked disorders and indicates that alternative approaches not initially based on patient cohorts are effective to reveal gene-disease associations. We provide a list of genes that are likely to be associated with human disorders. Further studies are required to delineate these disorders clinically and determine whether males and/or females harboring variants in these genes are affected.

## Data Availability

All data produced in the present work are contained in the manuscript or in the supplementary data.

https://www.genenames.org/

https://gnomad.broadinstitute.org/

https://gtexportal.org/home/

https://www.brainspan.org/

https://www.ensembl.org/index.html

https://genome.ucsc.edu/cgi-bin/hgTables

https://www.uniprot.org/

https://www.sysid.dbmr.unibe.ch/

https://www.ebi.ac.uk/gene2phenotype

## Supplementary Methods

### Protein-coding genes

Annotations of genes in all chromosomes were downloaded from the HUGO Gene Nomenclature Committee (HGNC) database in November 2020 focusing on protein-coding genes with approved status and present in the reference assembly.^51^ *MED14OS* was excluded from chrX-focused analysis due to its encoded protein being curated in Uniprot as “Product of a dubious CDS prediction”. Information concerning genes in PARs was also retrieved from HGNC.

### Gene-phenotype relationships

Gene associations with disorders and/or traits were retrieved from data files provided by Online Mendelian Inheritance in Man (OMIM). Information concerning the Clinical Synopsis of phenotypes was obtained through OMIM Search. OMIM morbid genes were annotated for 1) the availability of Clinical Synopsis data for any of the associated phenotypes, 2) the existence of Clinical Synopsis neurologic features in any of the associated phenotypes and 3) the presence of terms related to intellectual disability, seizures, impaired language development, impaired motor development, spasticity and ataxia among the Clinical Synopsis neurologic features. For the latter and due to the free-text nature of the Clinical Synopsis data, synonymous terms and sentences of the above-mentioned clinical features were used and their complete lists are shown in Supplementary Table 2. Genes were divided into three groups based on the existence and type of their gene-phenotype relationships: 1) “confirmed” when associated with at least one confirmed monogenic disorder; 2) “PMT” for genes either with provisional gene-phenotype relationship (P), or associated with susceptibility factors to multifacto-rial disorders (M) or with traits (T), which are labelled in the OMIM Gene Map with a question mark, braces or brackets, respectively; 3) “no disorder” for genes showing no phenotype associations. For each chromosome, we calculated the fraction of protein-coding genes that are CS-genes and the fraction of CS-genes that contain non-specific or specific neurologic features in the associated Clinical Synopsis data.

### Gene intolerance to variation

Metrics related to intolerance of chrX genes to genetic variation (loss-of-function (LoF), missense and synonymous) were retrieved from the Genome Aggregation Database (gnomAD) v2.1.1 ^23^ after conversion of gene identifiers to HGNC approved symbols. For genes with available data for multiple transcripts, the metrics of the one with lowest LoF observed/expected upper bound fraction (LOEUF) were kept.

### Gene expression data

Median gene-level transcripts per million (TPM) for 54 tissues from the v8 release were downloaded from the GTEx portal ^52^. The robust tissue specificity measure tau ^53,54^ was calculated for chrX genes as previously described, aggregating the median expression of each gene for the different brain regions into two values: i) the median of the values for the two cerebellar regions and ii) the median of the values for the other brain tissues.^22^ Genes with tau below 0.6 are broadly expressed, while tau higher than 0.6 indicates genes expressed in a restricted number of tissues.

Additional expression data was downloaded from the BrainSpan Atlas of the Developing Human Brain.^55^ The Developmental Transcriptome Dataset contains gene summarized RPKM (Reads Per Kilobase of transcript, per Million mapped reads) normalized expression values generated across 13 developmental stages in 8-16 brain structures from a total of 42 individuals of both sexes. After conversion of RPKM to TPM and gene identifiers to HGNC approved symbols, we restricted to genes on chrX and calculated the mean TPM according to age groups, ignoring sex and tissue origin: 1) Pre-natal; 2) Post-natal; 2a) Post-natal 1: after birth until four years-old (inclusive). 2b) Postnatal 2: older than four years old.

For the neural network, the expression data was stratified by sex: 1) we calculated the mean and variance expression of each gene for females and males independently, for brain, cerebellar tissues and nerve, aggregating in one metavalue the data for the two cerebellar tissues and in another the data for the other brain regions; 2) tau values were also calculated independently for females and males; 3) Brainspan mean and variance TPM for each age group was calculated separately for females and males.

### Canonical transcript selection

The criteria for canonical transcript selection prioritized: i) MANE (Matched Annotation between NCBI and EBI) Select transcripts, which were independently identified by both Ensembl and NCBI as the most biologically relevant; and ii) APPRIS-annotated transcripts ^56^. TSS, MANE, and APPRIS annotations of transcripts were retrieved from Ensembl Genes 102 via BioMart ^57^. For most genes (ca. 80%), the canonical transcripts belong to the MANE Select category. The longest transcript with APPRIS annotation was selected for ca. 20% of genes. In one case, the only existing transcript was kept.

### Promoter CpG density

CpG density was calculated for the 4kb region surrounding the transcription start site (TSS±2kb) of the canonical transcripts of chrX protein-coding genes following previous publications.^22,58^ Promoter sequences were downloaded through UCSC Table Browser.^59^ The CpG density, defined as the observed-to-expected CpG ratio, was calculated as follows: number of CpG dinucleotides / (number of C x number of G)) × N, where N is the total number of nucleotides in the sequence being analyzed.

### Exon and promoter conservation across species

Nucleotide conservation across 100 vertebrate species was calculated using the phastCons score obtained with the phastCons100way.UCSC.hg38 R package,^60^ and represents the probability that a given nucleotide is conserved (range 0 to 1). Exon-conservation score was calculated for each gene on chrX as the average phastCons of all nucleotides belonging to the gene coding sequence. Coding coordinates for the canonical transcripts were retrieved from Ensembl Genes 102 ^57^ using biomaRt R package.^61,62^ Promoter-conservation score for each gene on chrX was calculated as the average phastCons score of all nucleotides 4kb around the TSS of the canonical transcript.

### Distance to centromere and telomeres

The coordinates from centromeres and telomeres were downloaded through UCSC Table Browser. We calculated the distance from the TSS of the canonical transcripts to the centromere and to the telomere in the corresponding chromosomal arm.

### Annotation of encoded proteins

Data referring to function, subcellular location, subunit structure, and gene ontology terms for chrX encoded proteins were retrieved from Uniprot.^63^

### Paralogues

All paralogues from chrX genes were retrieve from Ensembl Genes 102 ^57^ using biomaRt R package.^61,62^ The 90^th^, 95^th^, 98^th^ and 99^th^ and 100^th^ percentiles were calculated for i) the percentage of paralogous sequence matching the query sequence and ii) the percentage of query sequence matching the paralogue sequence. Only paralogues with both metrics above the 95^th^ percentile were considered as close paralogues.

### X-chromosome inactivation

The information on genes escaping X-chromosome inactivation (XCI) was obtained from multiple publications.^4,64–69^ After converting gene identifiers from all studies into HGNC approved symbols, genes were divided into seven categories based on the agreement between the various studies: 1) high confidence escapee and 2) high confidence non-escapee, when almost all studies agreed on one status; 3) low confidence escapee and 4) low confidence non-escapee, whenever some studies disagreed but a higher number of studies reported one status; 5) variable escapee, when most studies stated variable escape; 6) discordant, when similar number of studies agreed on both status; 7) not available, when there was not enough data to have reliable evidence of XCI status.

### Features shared by disorder-causing genes

To uncover no-disorder genes exhibiting similar characteristics to known disorder-causing genes, we considered metrics showing enrichment of confirmed-disorder genes at one of the extremes of the distribution with marked difference between confirmed-disorder and no-disorder genes. Then, we applied a threshold approach to categorize genes showing values within the deciles enriched for confirmed disorder-causing genes for each of the considered metrics. The minimum number of criteria showing enrichment for confirmed disorder-causing genes was used as the minimum number of criteria required to consider a PMT or no-disorder gene as having similar features to disorder-causing genes.

### Pre-classification of genes for the neural network

Genes were pre-classified based on 1) the type of associations with disorders and/or traits (confirmed, PMT, no-disorder), 2) the association with a brain disorder and 3) the tolerance to loss-of-function (LoF) variants (Extended Data Fig. 5). The list of confidently LoF-tolerant genes based on the gnomAD dataset was retrieved from data files from Karczewski et al. (2020) and consists of genes with at least one homozygous LoF variants.^16^ Genes showing no homozygous LoF variants were considered LoF-intolerant.

### Neural Network

Information regarding the data fed into the neural network is in Supplementary Tables 7 and 8. We used a simple multilayer perceptron (MLP) network with 3 inner layers of size 128 nodes, 64 nodes and 32 nodes with rectified linear unit (ReLU) activation function and a single output node with sigmoid activation. The weights were initialized with a random normal distribution with a bias of zero, and batch normalization and a dropout of 0.5 were applied between all layers. Training was performed with a batch size of 5 and data shuffling between epochs using stochastic gradient descent (SGD) with a learning rate of 1e-3, no decay, and a binary crossentropy / log loss as the objective function.

Class labels for the training data *T* are “Cbi” (C1, value 1.0) and “NDt” (C0, value 0.0) (Extended Data Fig. 5; Supplementary Table 8). Beforehand columns of the training data were separately normalized to [0;1] via min-max-scaling.

To validate the model we performed a 10-fold cross validation by scikit-learn K-Fold ^70^, which randomly splits *T* into ten non overlapping parts *T*_0_ to *T*_9_ each containing 10% of the data (Extended Data Fig. 4). Ten networks are trained. Each network *N_i_* is trained on the nine parts {*P_j_*|*j* ≠ *i*}. Afterwards *N_i_* is applied to the remaining unseen part *P_i_* to create estimation *E_i_*.

We then concatenate all partial estimations to get full estimation ∪*_i_ E_i_* = *E* for each gene in *T*. Together with the known classes C0 and C1 for each gene, we estimate a threshold t for given false discovery rate d. First define functions *C*1(*t*) and *C*0(*t*) returning all genes of class C1 respectively C0 with estimation score < *t*. For *t* > 0.5 we can consider a gene with label T0 to be false positive. Thereby for a given *t* the empirical FDR can be measured by *T*1 (*t*)⁄(*T*0(*t*) + *T*1(*t*)). By calculating the FDR threshold by estimations on ten different neural networks, each applied on unseen data, we ensure that the threshold is not biased by overfitting. FDR thresholds were calculated as 0.8152034 (FDR 0.05) and 0.9745963 (FDR<0.01).

Finally, a full network *N* is trained on the full data *T* and applied on the complete data set with scaled column by the former min-max-scaling. The inferred score for each gene, represents the estimated probability for an association between the gene and a brain diseases.

All computational operations were performed in python with sklearn^70^ for normalization and keras for building and training the neural network.^71^ Seeds for numpy, K-Fold and Tensorflow were set to 1094795585.

### Known mutations

The number of indels, nonsense, splice site and missense variants reported in PMT and no-disorder genes were retrieved from the Human Gene Mutation Database (HGMD) Professional 2020.3 (Qiagen) and DECIPHER,^72^ and normalized by the CDS length. DECIPHER variants include DDD variants and patient sequence variants that are not benign or likely benign.

### Developmental disorders gene databases

chrX genes were annotated for their classification in SysID (https://www.sysid.dbmr.unibe.ch/)^73^ and Gene2Phenotype (https://www.ebi.ac.uk/gene2phenotype)^74^ developmental disorders databases, which list genes associated with intellectual disability or developmental disorders.

### Identification of damaging variants in predicted gene

*De novo* variants in predicted genes were retrieved from Kaplanis et al.^38^ and Martin et al.^21^ In addition, we examined variants in predicted genes identified in unsolved cases with developmental disorders from two different cohorts: a cohort from Hôpital Pitié-Salpêtrière (APHP, Paris, France), which mainly includes patients with intellectual disability or neurological disorders (6500 individuals from 2346 families) and a cohort of patients with dysgenesis of the corpus callosum, associated with learning or intellectual disabilities from University of California San Francisco (UCSF; 1399 individuals from 463 families). Informed consent of the legal representatives and appropriate approval of an ethics committee, according to the French and American laws, have been obtained. Variant data and clinical information were shared anonymously. Only damaging variants (*i.e.* variants leading to a premature termination codon or missense variants with CADD PHRED scores ≥ 25) in NN-predicted genes meeting at least one LME criterion were kept for further analysis.

### Functional validation of *TRPC5* missense variant

HEK293 cells were transiently transfected with a plasmid expressing TRPC5-GFP WT or p.Arg175Cys mutant channels. Currents were recorded from green fluorescent cells using the whole-cell configuration of the patch-clamp technique 16 to 24 h after transfection at room temperature. Voltage-clamp recordings were performed using an Axopatch 200B amplifier and a Digidata 1440 Analogue/Digital interface (Axon Instruments). Data were low-pass filtered at 2 kHz, digitized at 10 kHz and analysed offline using Clampfit software. Currents were recorded during a 500 ms voltage ramp from –100 mV to 100 mV applied from a holding potential of –100 mV every 5 s. Series resistance was not compensated, and no leak subtraction was performed. Data were not corrected offline for voltage error and liquid junction potential. The pipette solution contained (in mM): NaCl 8, Cs-methanesulfonate 120, MgCl2 1, CaCl2 3.1, EGTA 10, HEPES 10, pH adjusted to 7.3 with CsOH. The extracellular solution contained (in mM): NaCl 140, MgCl2 1, CaCl2 2, HEPES 10, glucose 10, pH adjusted to 7.2 with NaOH. Immediate currents were recorded upon break-in (using patch pipettes that contained 100 nM free Ca2+). The mutant currents are readily distinguished even in the absence of agonist stimulation, indeed the current-voltage relationship (IV) was similar to that described in the literature, showing inward and outward (’double’) rectification, giving something that is roughly ’Nshaped’. Englerin A (100nM) was applied to the cell expressing WT and mutant TRPC5 without immediate current upon break-in to confirm its functional expression.

### Statistics

Fisher‘s tests were performed for each chromosome to determine associations between: i) genes being present in the specific chromosome and genes having an associated phenotype with Clinical Synopsis data (CS-genes); ii) CS-genes being in the specific chromosome and CS-genes having non-specific or specific neurologic features described in the Clinical Synopsis data. For chrX, Fisher’s tests were performed to compute the enrichment/depletion of confirmed disorder-associated genes in each decile of the distribution of continuous variables. Odds ratios were log2 transformed and indicate enrichment or depletion of genes, for positive or negative values, respectively (10^-4^ was added prior to transformation, whenever necessary to deal with log of 0 issue). *P*-values were adjusted for multiple comparisons using Bonferroni correction. For gene prediction, Fisher’s tests were performed to calculate the association between genes being confirmed-disorder genes and the number of predictors. Mann-Whitney U test followed by Bonferroni correction for multiple testing was used to assess the coding-sequence length difference between groups of genes.

## Data availability

All data are available in the main text or supplementary materials. We used data from HGNC (https://www.genenames.org/), OMIM (https://www.omim.org/), gnomAD v2.1.1 (https://gnomad.broadinstitute.org/), GTEx Portal (https://gtexportal.org/home/), BrainSpan Atlas of the Developing Human Brain (https://www.brainspan.org/), Ensembl (https://www.ensembl.org/index.html), UCSC Table Browser (https://genome.ucsc.edu/cgibin/hgTables), Uniprot (https://www.uniprot.org/), Human Gene Mutation Database (http://www.hgmd.cf.ac.uk/ac/index.php), DECIPHER (https://www.deciphergenomics.org/), SysID database (https://www.sysid.dbmr.unibe.ch/), Gene2Phenotype (https://www.ebi.ac.uk/gene2phenotype) and CADD - Combined Annotation Dependent Depletion (https://cadd.gs.washington.edu/).

## Code availability

The following publicly available software packages were used to perform analyses: biomaRt (https://bioconductor.org/packages/release/bioc/html/biomaRt.html), phastCons100way.UCSC.hg38 (http://bioconductor.org/packages/release/data/annota-tion/html/phastCons100way.UCSC.hg38.html), rstatix (https://cran.r-project.org/web/packages/rstatix/index.html), numpy (https://numpy.org/), scikit-learn K-Fold (https://github.com/scikit-learn/scikit-learn) and Tensorflow (https://www.tensorflow.org/).

## Acknowledgements

The authors thank the patients with *CDK16* and *TRPC5* pathogenic variants and their family for their participation in this study. We kindly thank Mrs Céline Cuny for Sanger sequencing analysis of the patient with *TRPC5* missense variant. Electrophysiological experiments were carried out at the electrophysiology core facility of ICM funded from the program “Investissements d’avenir” ANR-10-IAIHU-06. We thank Universitätsklinikum Essen, University Duisburg-Essen, the Deutsche Forschungsgemeinschaft (DFG), the Tom-Wahlig-Stiftung (TWS), the Deutsche Stiftung Neurologie (DSN), and Assistance Publique des Hôpitaux de Paris (APHP) for their financial support to the research studies conducted by the authors. E.L. and F.K. are associated with the FOR 2488 project (DFG, Project number 287074911). This study makes use of data generated by the DECIPHER community. A full list of centres who contributed to the generation of the data is available from https://deciphergenomics.org/about/stats and via email from. Funding for the DECIPHER project was provided by Wellcome.

## Author Contributions

E.L. and C.De. conceived and supervised the study. E.L. and C.S. performed the computations. E.L., I.P., C.Da., A.R., T.K., S.K., N.D., and C.E. carried out the experiments. A.K., B.G., E.S., C.N., J.P., B.D-B., L.V., A.P.A.S., E.K.V., J.A.J.V., F.J.K., F.T.M-T., M.S., P.S., S.G.M.F., E.A., E.H.S., F.E., J.B., B.K., C.M., D.H., J-L.M., J.G., V.M.K., B.H., and A.P. contributed analysis, data and/or critically revised the manuscript for intellectual content. E.L. and C.De. drafted the manuscript. All authors reviewed and approved the manuscript.

