## Extended Data for "Systematic analysis and prediction of genes associated with disorders on chromosome X"

**Extended Data Fig. 1: a-b**, Genes associated with motor impairment, spasticity or ataxia are not enriched nor depleted in specific chromosomes. **a,** Per chromosome enrichment/depletion of CS-genes having specific neurologic features described in the Clinical Synopsis data (Fisher's test followed by Bonferroni correction for multiple testing). **b,** Fraction of CS-genes that contain specific neurologic terms in the associated Clinical Synopsis data. **c-d,** Comparisons of morbid genes with Clinical Synopsis information between chromosomes show similar results. **c,** Per chromosome enrichment/depletion of i) protein-coding genes in morbid genes with at least one associated phenotype comprising Clinical Synopsis data (morbid CS-genes) (first graph) and ii) morbid CS-genes in genes with non-specific or specific neurologic features described in the Clinical Synopsis data (other graphs). Only significant p-values are shown (Fisher's test followed by Bonferroni correction for multiple testing). **d,** Fraction of i) morbid CS-genes among protein-coding genes or ii) morbid CS-genes that contain non-specific or specific neurologic terms in the associated Clinical Synopsis data. **a-d,** Yellow, enrichment; dark-grey, depletion; light-grey, not significant. Specific neurologic terms: intellectual disability, seizures, language impairment, motor impairment, spasticity and ataxia. Synonymous terms/sentences were used in OMIM searches (Supplementary Table 2).

**Extended Data Fig. 2: Analyses of continuous variables distributions. a,** Barplots showing the enrichment of confirmed genes in each decile of continuous variables distributions. Yellow, enrichment; dark-grey, depletion; light-grey, not significant. Only significant p-values are shown (Fisher's test followed by Bonferroni correction for multiple testing). **b,** Density plots showing the distribution of continuous variables according to gene group. Genes associated with at least one monogenic disorder (confirmed genes, orange), genes with provisional associations or associated with susceptibility factors to multifactorial disorders or with traits (PMTs, blue), and genes without associated phenotypes (no-disorder genes, grey). Vertical dashed lines separate deciles of the overall distribution. Grey areas depict deciles for which confirmed disease-causing genes are enriched (related to panel a).

**Extended Data Fig. 3: a-d,** Paralogues of protein-coding genes on chrX. Density plots showing the distribution of target (**a and b**) and query (**c and d**) percentage of identity for all paralogues of protein-coding genes on chrX according to gene group (**a and c**) or predicted status after the NN approach (**b and d**). Vertical dashed lines depict the 95th percentile of the overall distribution (81.4 and 83.9% for target and query percentages of identity, respectively), and grey areas mark paralogues above the 95th percentiles.
**e,** Boxplot showing a bias of LOEUF and misZ criteria against genes with smaller coding-sequence (CDS) length. Box plot elements are defined as follows: center line: median; box limits: upper and lower quartiles; whiskers: 1.5× interquartile range; points: outliers. **f,** Enrichment of confirmed genes in genes meeting at least one of the L, M or E criteria. Yellow, enrichment; dark-grey, depletion; light-grey, not significant. Significant p-values are shown (Fisher's test followed by Bonferroni correction for multiple testing).

**Extended Data Fig. 4: Overview of the neural network. a,** Layers and nodes used in the neural network. ReLU, Rectified Linear Unit. **b,** Visualization of the training, quality control by 10-fold cross validation and estimation of the false discovery rate of the neural network. The training data is randomly divided into ten different parts of equal size. Ten networks (purple centered squares) are trained independently on nine different data blocks. The tenth part is used for prediction (example shown for network 5). All predictions are combined into an overall prediction (upper blue rectangle) and used to estimate a threshold for a given FDR by the algorithm described in this work. The final model (lower purple rectangle) is trained with the unsplit training and applied to the unknown data (lower left) to obtain a final prediction for each gene.

**Extended Data Fig. 5:** Neural network predicts putative disorder genes. **a,** Protein-coding genes were pre-classified into 10 subgroups based on 1) the type of associations with disorders and/or traits (confirmed, PMT, no-disorder), 2) the association with a brain disorder and 3) the tolerance to loss-of-function (LoF). Two classes were used to train the neural network: Cbi (confirmed brain-disorder associated genes that are LoF-intolerant; value 1.0) and NDt (no-disorder genes tolerant to LoF mutations; value 0). We show number and fraction of predicted genes (FDR<0.05) for each of the 10 classes for chrX and autosomes separately. **b,** Empirical cumulative distribution functions of the prediction value for each of the 10 classes. Region corresponding to FDR threshold is shaded in grey. **c,** Per chromosome fraction of predicted disorder genes intolerant to LoF variants and associated with neurological features (Cbi). **d,** Fraction of predicted Cbi genes for each of the 10 subgroups, in chrX (black) and in autosomes (grey). **e and f,** Per chromosome enrichment/depletion of Cbi genes (**e**) and correctly predicted Cbi genes at FDR<0.05 (**f**). Fisher's test followed by Bonferroni correction for multiple testing.

**Extended Data Fig. 6: Known point mutations in no-disorder genes. a-b,** Scatter plots showing the correlation between the coding-sequence (CDS) size and the number of known HGMD (**a**) and DECIPHER (**b**) mutations. **c-d**, Boxplot showing the number of known mutations reported on HGMD (**c**) and DECIPHER (**d**) for no-disorder genes according to their predicted status. **e-f**, Boxplot showing the number of known mutations normalized by coding-sequence (CDS) length due to the small correlation between the two variables that were reported on HGMD (**e**) and DECIPHER (**f**) for no-disorder genes according to their predicted status. **c-f,** Box plot elements are defined as follows: center line: median; box limits: upper and lower quartiles; whiskers: 1.5× interquartile range; points: outliers. Related to Supplementary Table 9.

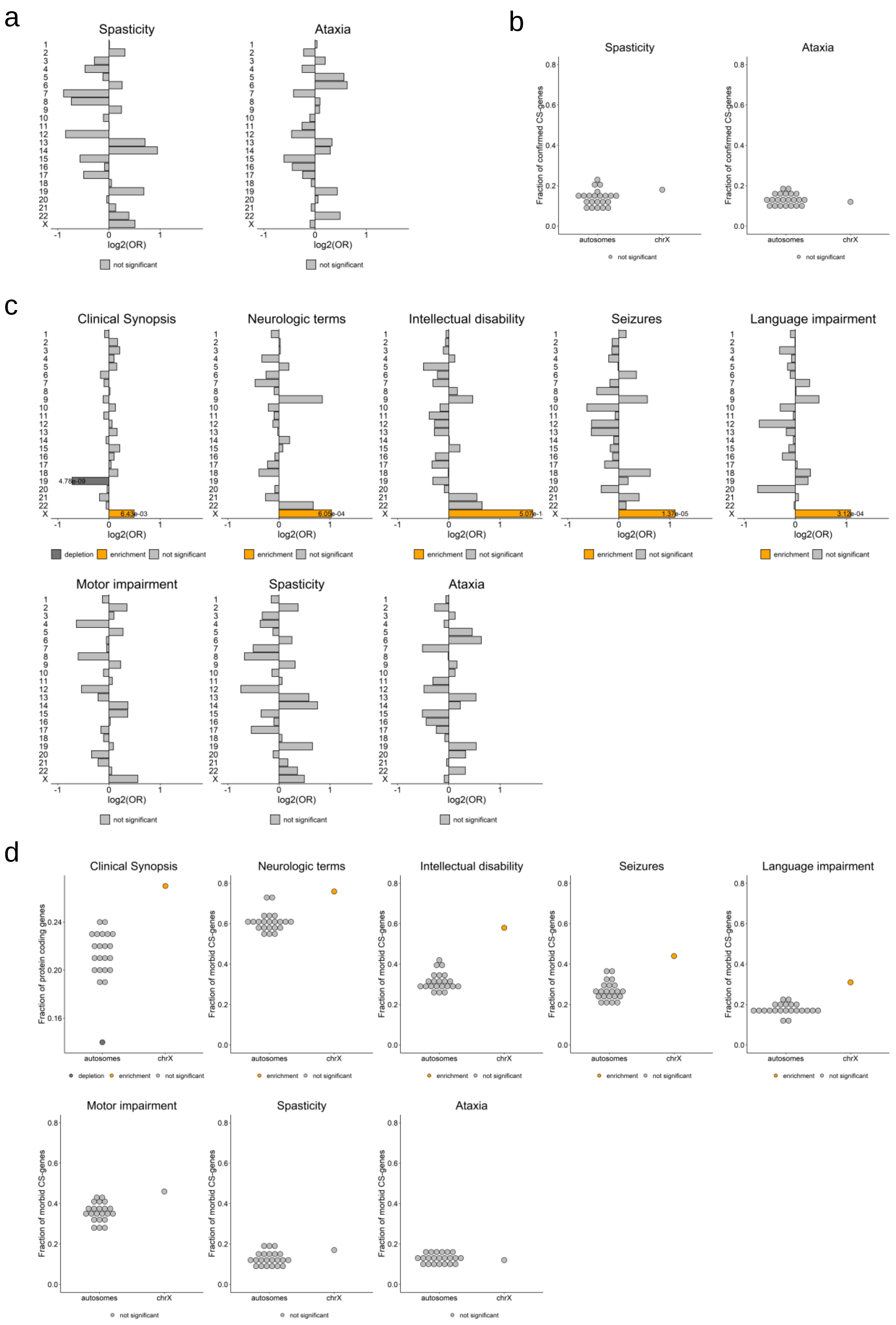


**Extended Data Fig. 1**

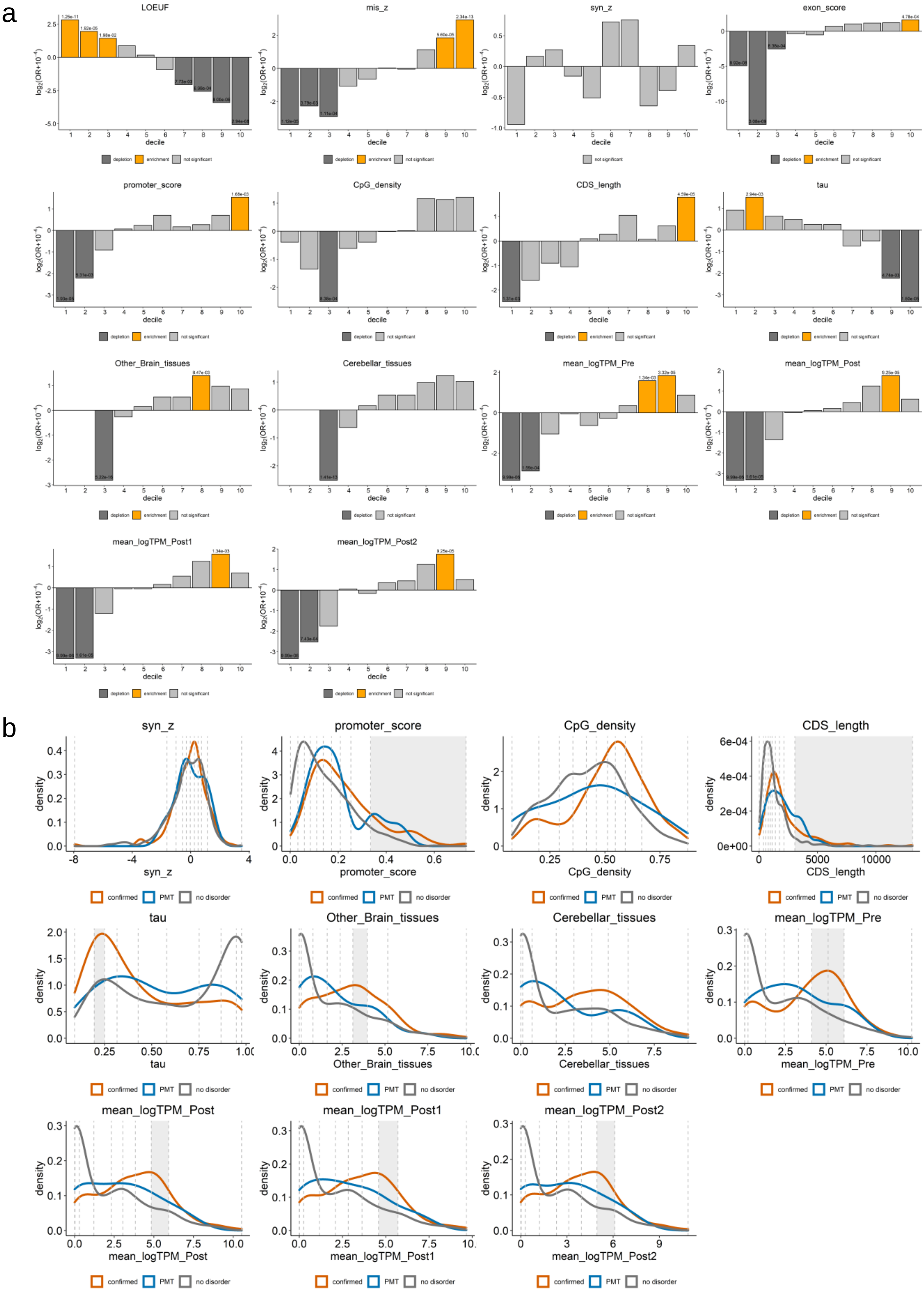


**Extended Data Fig. 2**

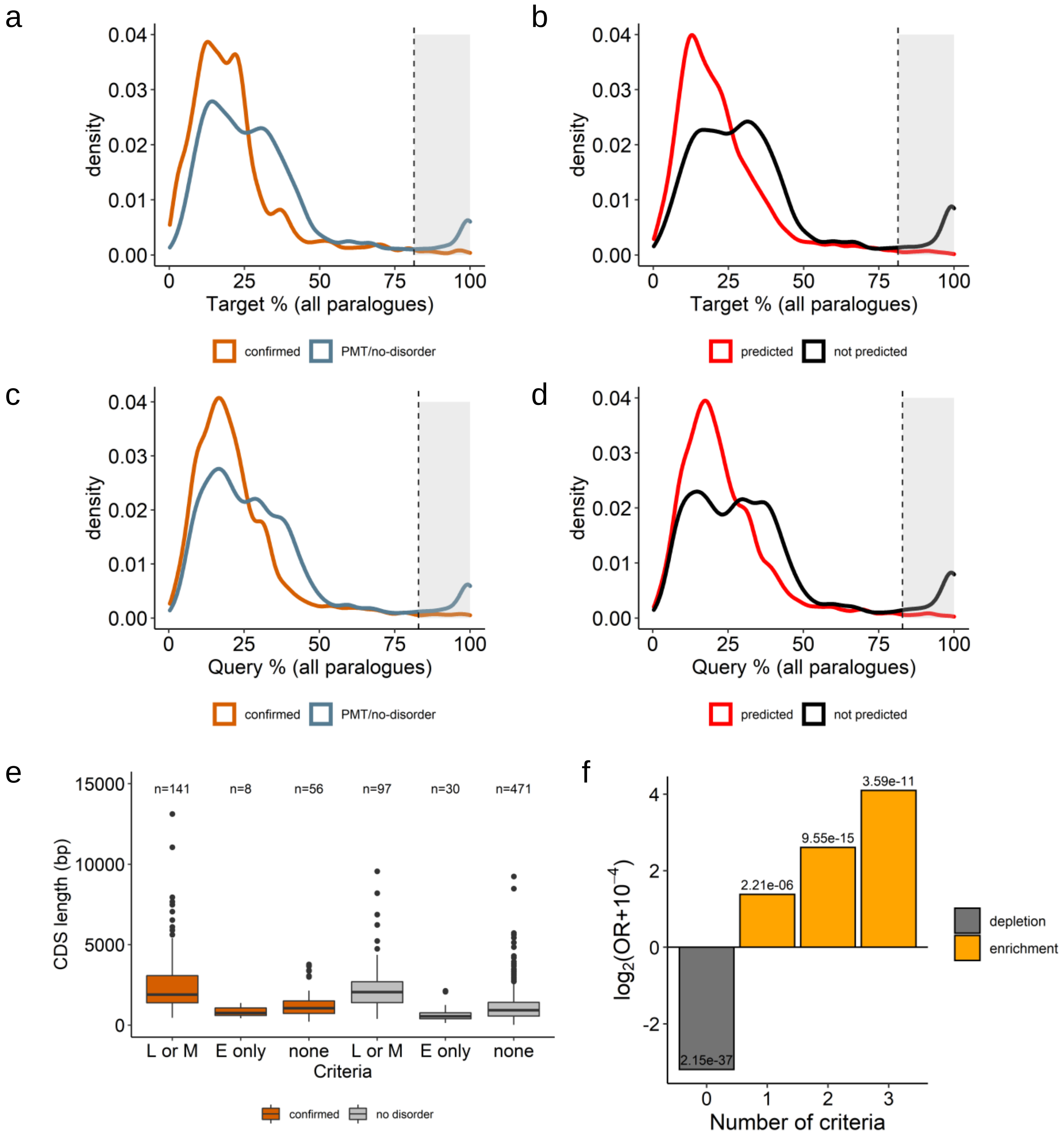


**Extended Data Fig. 3**

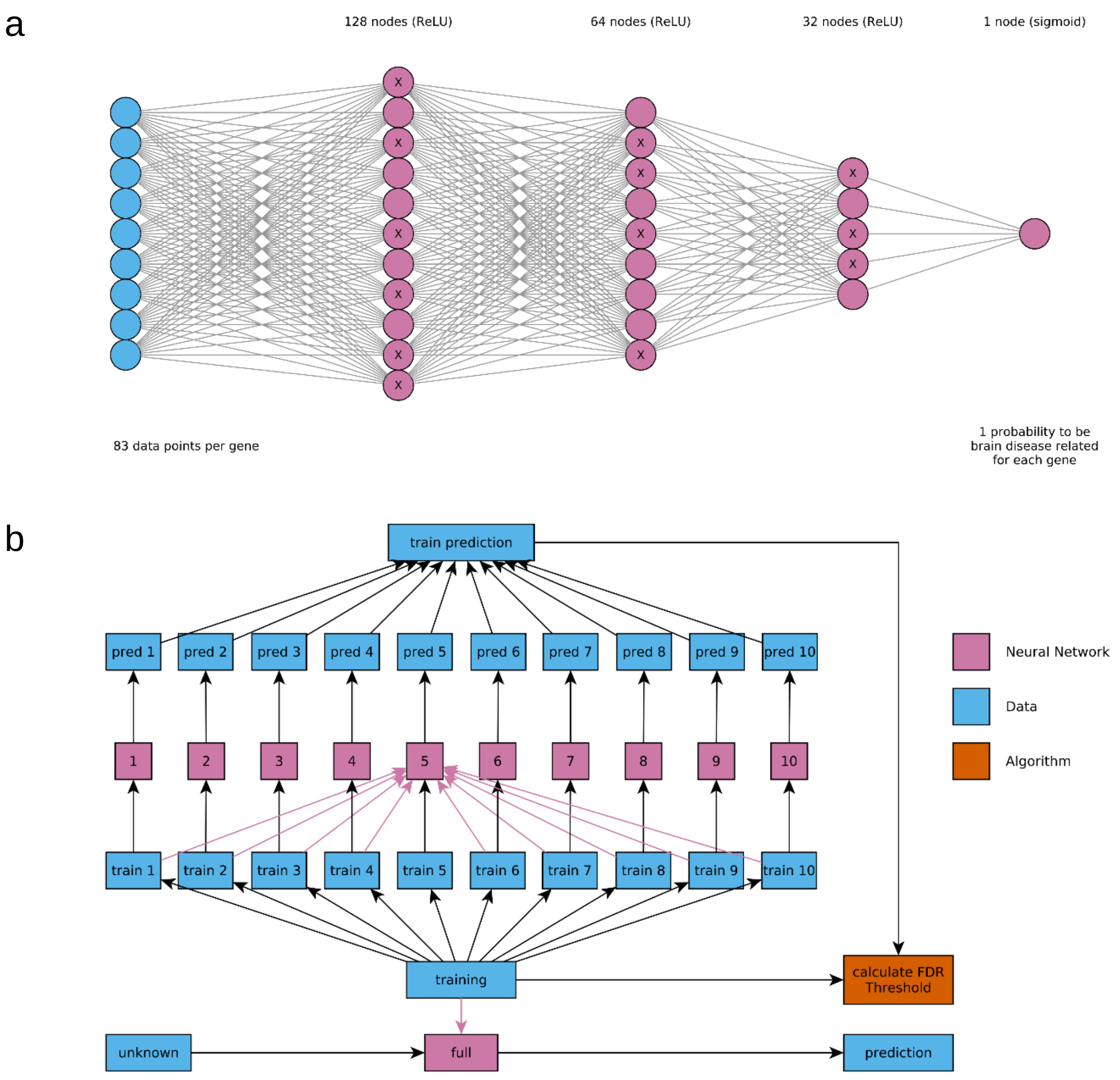


**Extended Data Fig. 4**

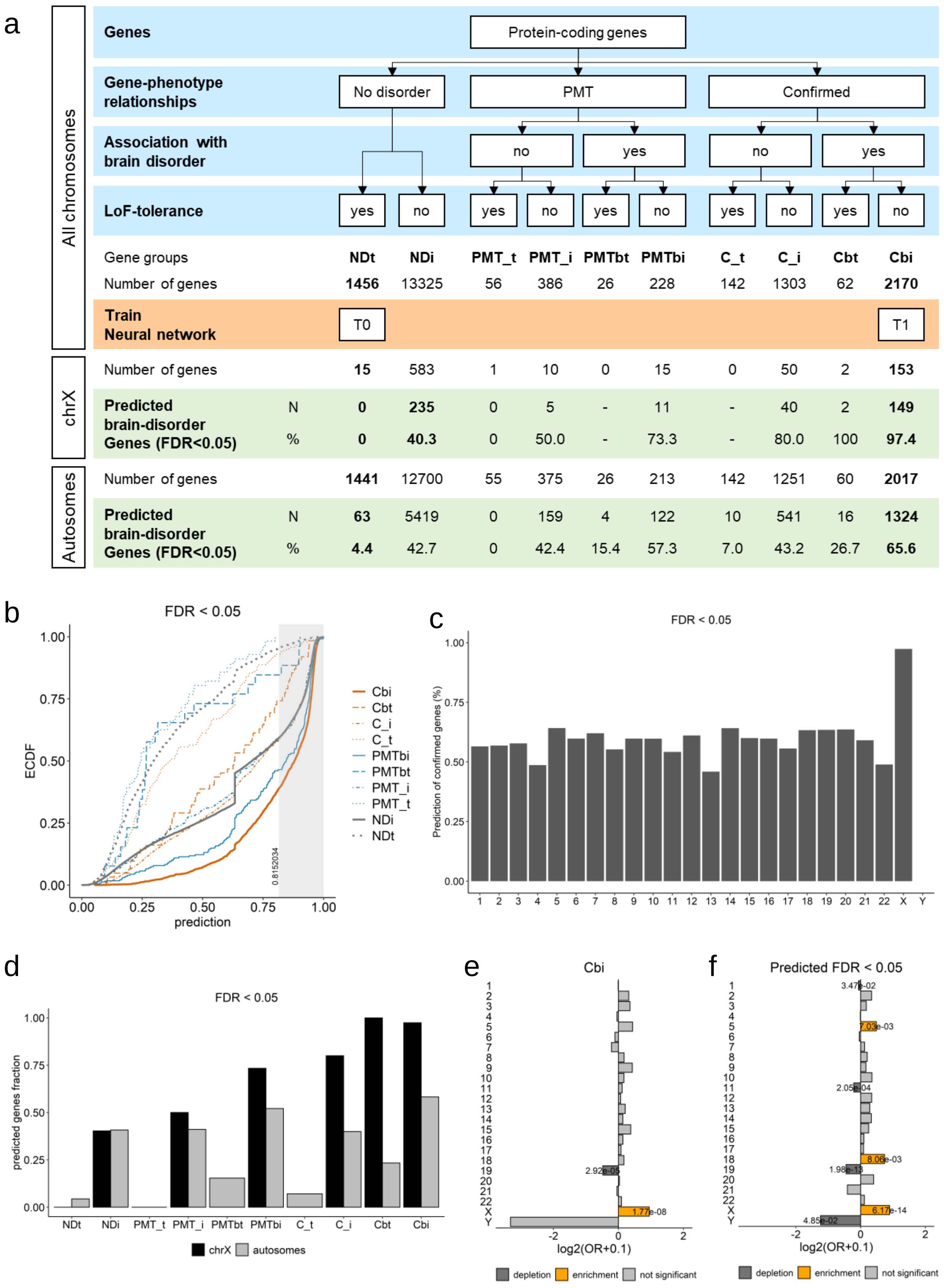


**Extended Data Fig. 5**

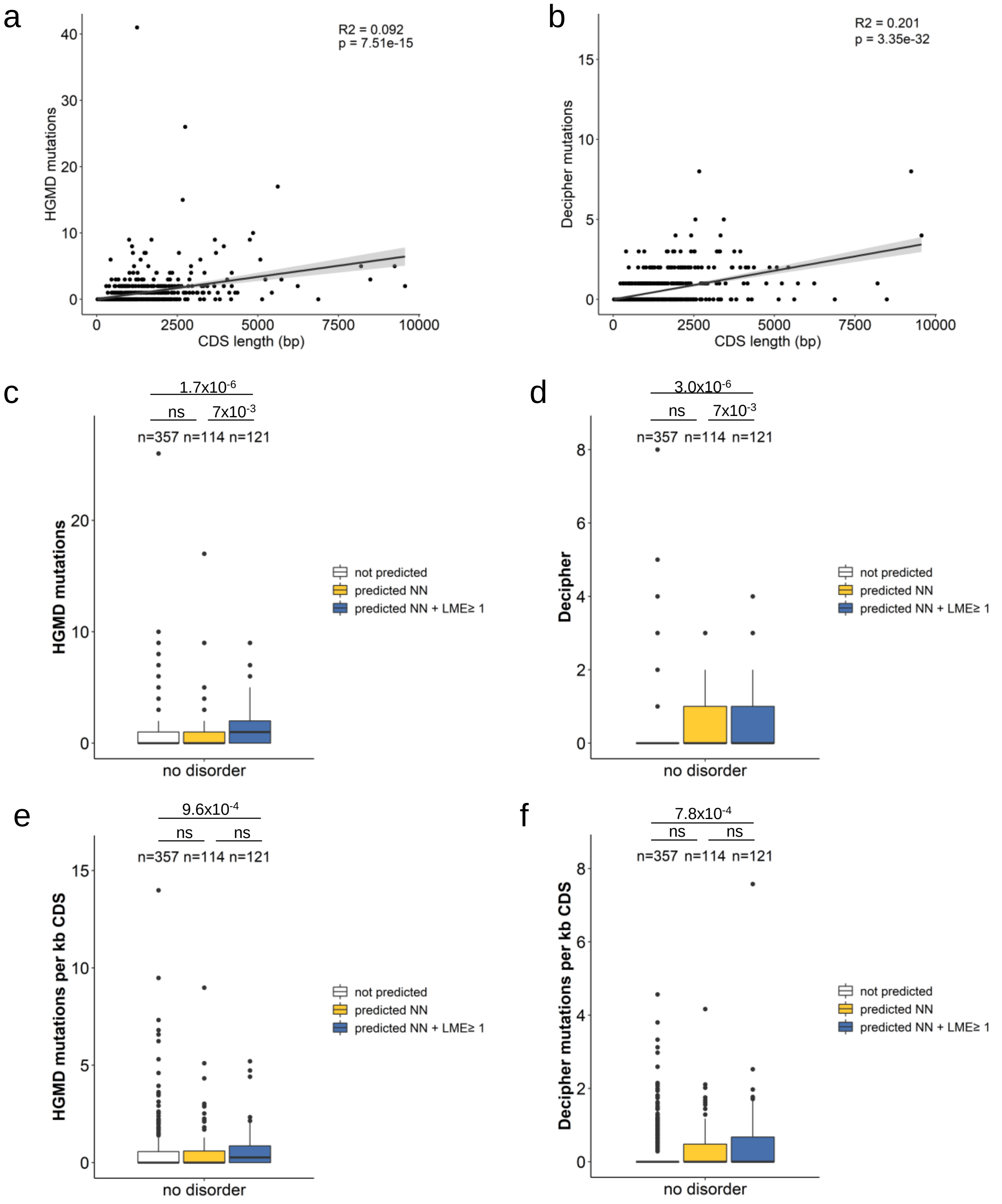


**Extended Data Fig. 6**
